# Over- and under-estimation of vaccine effectiveness

**DOI:** 10.1101/2022.01.24.22269737

**Authors:** Hilla De-Leon, Dvir Aran

## Abstract

**Background:** The effectiveness of SARS-CoV-2 vaccines has been a subject of debate, with varying results reported in different studies, ranging from 60-95% vaccine effectiveness (VE). This range is striking when comparing two studies conducted in Israel at the same time, as one study reported VE of 90-95%, while the other study reported only ~80%. We argue that this variability is due to inadequate accounting for indirect protection provided by vaccines, which can block further transmission of the virus

**Materials and Methods:** We developed a novel analytic heterogenous infection model and extended our agent-based model of disease spread to allow for heterogenous interactions between vaccinated and unvaccinated across close-contacts and regions. We applied these models on real-world regional data from Israel from early 2021 to estimate VE using two common study designs: population-based and secondary infections

**Results:** Our results show that the estimated VE of a vaccine with efficacy of 85% can range from 70-95% depending on the interactions between vaccinated and unvaccinated individuals. Since different study designs capture different levels of interactions, we suggest that this interference explains the variability across studies. Finally, we propose a methodology for more accurate estimation without knowledge of interactions

**Discussions and Conclusions:** Our study highlights the importance of considering indirect protection when estimating vaccine effectiveness, explains how different study designs may report biased estimations, and propose a method to overcome this bias. We hope that our models will lead to more accurate understanding of the impact of vaccinations and inform public health policy.

## INTRODUCTION

Hundreds of millions of individuals have been infected with the SARS-CoV-2 virus, and millions of them died because of the infection since it was first detected in China at the end of 2019. In response, several companies and research institutions developed vaccines to help suppress the pandemic, and less than a year after identifying of the virus, several clinical trials reported results of proven vaccine efficacy (1,2). The randomized clinical trials of the Pfizer/BioNTech and Moderna vaccines showed vaccine efficacy of around 95% in the first months after the second dose; however, the studies had relatively low numbers of infected individuals, and this level of efficacy can only be regarded as a proxy to the real-world vaccine effectiveness across divergent populations, demographics, and clinical features.

By the end of 2020, many countries launched mass vaccination campaigns. The data collected through these mass vaccination campaigns was used to evaluate the effectiveness of the vaccines in a wide range of populations (3–15). Individual-level study designs for vaccine effectiveness evaluation can be split into two main approaches: (1) A population-based approach (PB), where the number of vaccinated and unvaccinated infections are counted after correcting for possible confounders (3); (2) A secondary infections approach (SI), which allows to overcome a major limitation of the first approach of unknown exposure. In this approach, infections are counted after a known exposure, such as secondary household infections (4). Interestingly, when examining the vaccine effectiveness estimations from studies that use each approach, there are major differences. In PB studies, vaccine effectiveness was usually estimated at 90%-95% (1–3,5), while in SI studies, much lower vaccine effectiveness estimations are usually reported (60%-80%) (4,11,12). Moreover, observational studies in nursing home residents reported varying vaccine effectiveness levels ranging from 53% to 92% against SARS-CoV-2 infection (6,13,14). The difference between the two effectiveness levels is emphasized when we compare the 90%-95% effectiveness of Dagan et al. (3) with the 80% effectiveness of Gazit et al. (4). Both studies evaluated the effectiveness of the same vaccine in a similar population in Israel at the same time, and only the study design differed. We hypothesize that the main reason for this gap can be explained by the proportion of interactions of vaccinated individuals with other vaccinated individuals that diverges between the two settings. Our hypothesis is based on one feature of the vaccines that has been broadly neglected in current vaccine effectiveness studies. While the vaccines were designed mainly to reduce symptomatic disease, severe disease, and death, studies from 2021 have shown that they also provide efficient transmission-blocking. A study from Sweden found that the number of immune members in a family was negatively correlated with the likelihood of incidence of infection of non-immune family members (16). Similarly, a study from Israel found reduced infection rates in communities as vaccination rates increased (17). Previous studies before COVID-19 have shown how interference, the potential outcomes of one individual are affected by the treatment assignment of other individuals, can affect vaccine effectiveness estimations (18). It seems that an underlying assumption made by the COVID-19 vaccine effectiveness studies is that there is no interference, which is an unfounded assumption.

Previous studies presented causal inference frameworks with interference to deal with the effects of indirect protection on vaccine effectiveness estimations (18–23). The main novelty of this work stems from our attempt to use a numerical model that incorporates real-world data on vaccination and infection rates to demonstrate how different study designs may lead to divergent estimations of VE. Here, we developed both an analytical approach and a numerical approach which utilizes our Monte Carlo Agent-based Model (MAM) (24) to examine the influence of interference between vaccinated and unvaccinated individuals on data of COVID-19 vaccinations and infections in Israel (25,26). Our analyses show that different assumptions may lead to significant under- or over-estimations and might explain the differences in estimations between the different study design approaches. We suggest that this divergence is inversely related to the rate of mixing between vaccinated and unvaccinated individuals, in line with previous studies on interference (19,20,23). Since it is not simple to estimate the level of mixing, which diverges across time and communities, we developed a simple approach that can overcome this issue and provide accurate estimations of vaccine effectiveness regardless of mixing levels.

## MATERIALS and METHODS

The main aim of this paper is to quantify vaccine effectiveness estimations in terms of interference between vaccinated and unvaccinated individuals. Interference occurs when the treatment received by other individuals influences the effects of an exposure or intervention on an individual. Because interference can affect the relationship between exposure and outcome, it can be harder to determine causality. In terms of this work, we define interference as the effect on the risk of infection for a vaccinated individual due to exposure to an unvaccinated individual.

All the data and code for modelling and figures presented in this manuscript are available on: https://github.com/hdeleon1/Over-and-under-estimation-

### Analytical infection model

First, we developed a simple heterogenous analytical infection model to find a quantifiable relationship between the amount of interference in a network and the effectiveness of vaccines (see **Supplementary Methods**; **Table S1-2** for more information). Within the model, we define a network of vaccinated and unvaccinated nodes. Each node is strongly connected to all the (*N*_*in*_ − 1) nodes in its clique, and weakly contacted to all *N*_*out*_ other nodes (*N*_*all*_ *= N*_*in*_ *+ N*_*out*_) with *δ* as the ratio between the number of infections that originate from the inner clique and the other infections.

Using the model we can define the observed vaccine effectiveness, 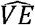, as a function of the average fraction of edges with unvaccinated nodes to the vaccinated nodes, ⟨*I*⟩:

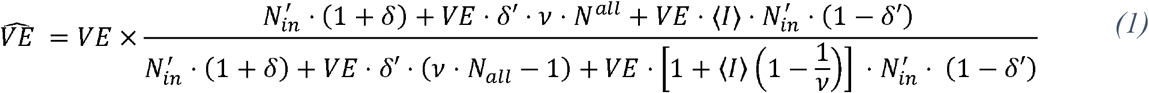

With *v* as the fraction of unvaccinated people in the total population, 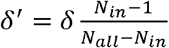 and 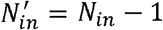.

To summarize, this analytical model is a relatively simple model that simulates infections in a population with non-uniform vaccination rates and at heterogenous circuits of infection risks. It considers the risk of infection in vaccinated households and allows to quantify the population 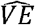, corrected by the interference, as opposed to the individual-level vaccine effectiveness.

### MAM – A flexible Monte-Carlo Agent based Model

To model the infections occurred in the real-world we use MAM, a flexible Monte-Carlo Agent based Model for modelling COVID-19 spread that accurately predicts infection dynamics in Israel (24). In this work, we expand this model to include complex interaction networks, such as differing between household interactions, which represent around half of infections in Israel (27), and other interactions. The spatial feature of MAM allows it to be used to predict both the local and global spread of COVID-19 by simultaneously modeling spatial interactions between families (high interaction), neighborhoods (medium interaction), and cities (low interaction). By applying MAM to public data from Israel, we could model the actual spread of COVID-19 during Israel’s first vaccination campaign for 9.2 million particles across 1,578 separate communities, where each community has its own vaccination rate.

## RESULTS

Most transmissions of the virus leading to infection are in close interactions. These interactions are often cliques, where all individuals in the clique have close interaction with each other (i.e., households). Heterogenous vaccination rates between those cliques bias the estimations of vaccine effectiveness. Consider this simple example: two nursing homes, each with 60 people that all interact with each other. Assuming that in one home 50 people are vaccinated with a vaccine with 80% efficacy and the rest are unvaccinated, while in the other it is the opposite (10 vaccinated). Both homes had an outbreak, in the first house one vaccinated and one unvaccinated were infected, which is what we expect with 80% protection. In the second home 25 unvaccinated and one vaccinated were infected, again translates to 80% protection. However, if we calculate 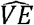 for both homes together (120 people in total), we substantially overestimate 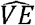 at 92.3%, which is 2.6-fold risk reduction. Similar analysis can be performed with secondary infection models to show any desired level of observed 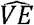. We present here an analytical framework to model this observation.

### Analytical infection model

For illustration purposes, we present three possible mixtures described using three networks. In all networks, there is a similar amount of vaccinated and unvaccinated nodes, and there are precisely four close connections for each node (*N*_*in*_ *=* 5, *N*_*out*_ *=* 20) (**Figure 1A-C**). In each network all the nodes are numbered, such that for the *i*^*th*^ node we indicate with vector *A*^*i*^ its vaccination status and the number of connections it has with the unvaccinated nodes in the network (see **Supplementary Methods**). The only difference between the three networks is the dispersal of the vaccinated and unvaccinated nodes: in network (A), there is complete segregation between vaccinated and unvaccinated nodes (⟨*I*⟩ = 0). This network might simulate nursing homes that were vaccinated early, while most of the rest of the population was not yet vaccinated. In network (B), for each vaccinated node, half of the edges are to vaccinated nodes and half to unvaccinated nodes (⟨*I*⟩ = 0.5). In network (C), each vaccinated node is connected to one vaccinated node and three unvaccinated nodes (⟨*I*⟩ = 0.75). Network (C) might simulate households where two members are vaccinated (e.g., parents) and three are not, e.g., children. The *A*^*i*^ values for each node in the networks are provided in the **Supplementary Methods** (**Table S3**).

**Figure 1.**
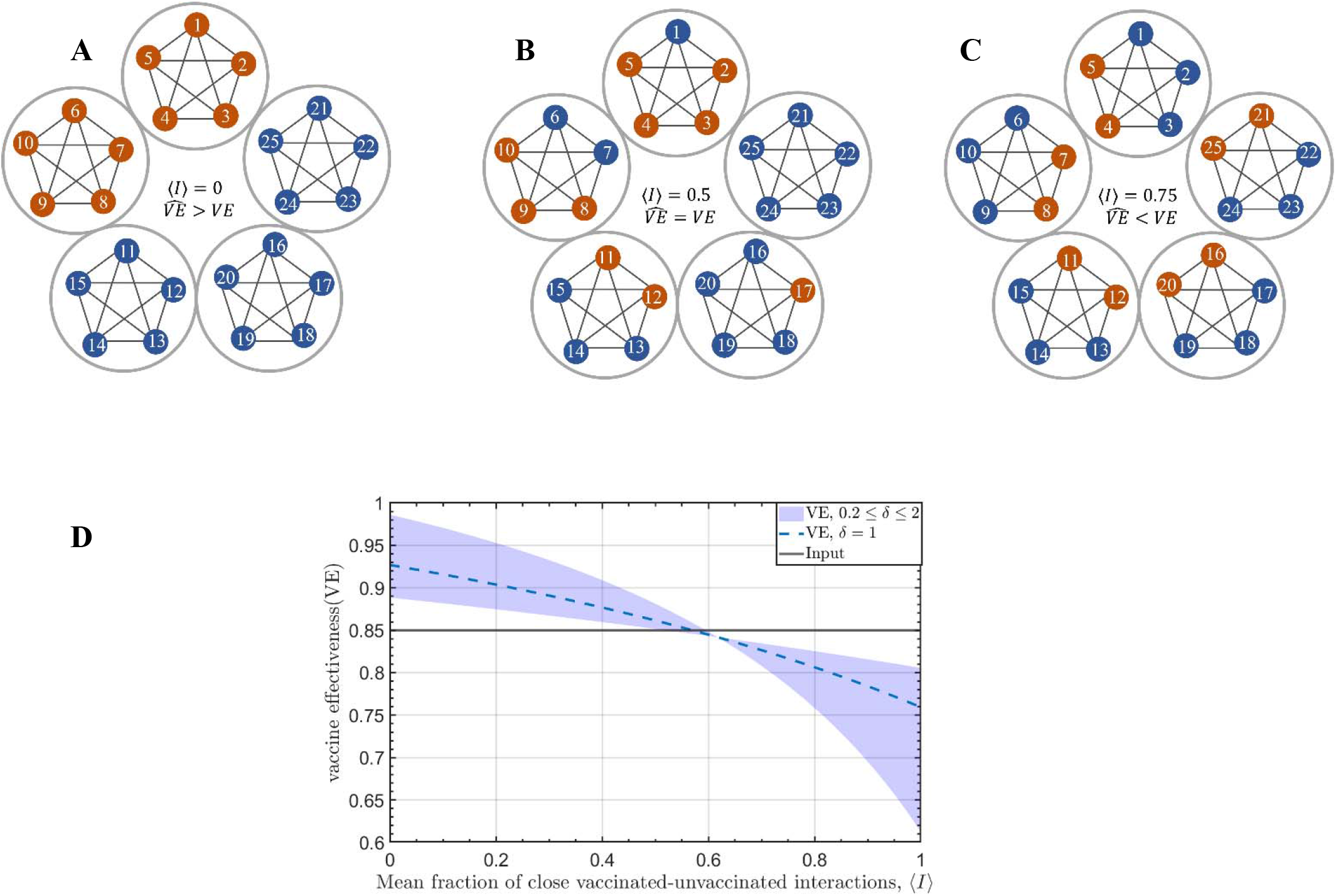
Infection networks with different types of interactions between vaccinated and unvaccinated nodes. **A-C**. Orange individuals are vaccinated nodes, blue nodes are unvaccinated nodes. Each circle is a tightly connected clique. Weak interactions with individuals outside of the clique. **D**. 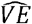 as a function of⟨I⟩, for δ = 1, and vE = 0.85: Blue band: 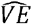 for 0.2 < δ < 2; Blue dashed line: **δ = 1**; Solid line: the input vE.

For each of the networks we count the average fraction of edges with unvaccinated nodes to the vaccinated nodes, ⟨*I*⟩, and calculate 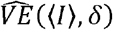 for *VE* = 0.85 and *v* = 0.6 (proportion of the population that are unvaccinated), and *δ* = 1 (ratio between inner clique and other infections, thus 1 represents that half of the infections are household infections). Using the equations described in the analytical infection model, we find that the risk of infection in (B) for vaccinated nodes is 0.072 and 0.52 for unvaccinated nodes, which translates to 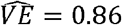, like *VE*. However, in (A) and (C) we see a diversion from *VE*: in (A) risk for vaccinated is 0.046 and 0.63 for unvaccinated, which translates to an overestimation of almost double reduced risk 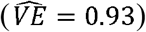, and in (C) these numbers are 0.1 and 0.4, respectively, which translate to an underestimation of 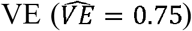 Generally, we observed that for ⟨*I*⟩ around 0.6 *VE* is similar to 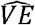 (**Figure 1D** and Eq (S-18) in the **Supplementary Methods**); **Figure 1D** shows 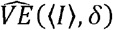 for 0 ≤ ⟨*I*⟩ ≤ 1 and 0.2 ≥ *δ* ≥2. Lower ⟨*I*⟩ levels produce overestimations of 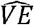 up to 95%, and higher ⟨*I*⟩ levels produce underestimations towards 70%.

### Numerical analysis: using MAM to estimate vaccine effectiveness in Israel

We next attempted to model the infections occurred in real-world and show how calculations of vaccine effectiveness can be skewed based on different assumptions regarding ⟨*I*⟩. Using MAM, we simulated the spread of COVID-19 infections in Israel from the beginning of the vaccination campaign on December 20, 2020. The particles were numbered and represented by the vector *A*^*i*^, similarly to the analytical model. However, since this is a time-dependent model, and the model predicts the spread of infection while the population is actively being vaccinated, we use *A*^*i*^(*t*) where the vaccination rate is determined by the actual vaccination rates across 1,578 communities in Israel (25,26) (**Figure S1**). We created ten different mixing scenarios between vaccinated and unvaccinated individuals in each community. For each scenario, we created a different array of *A*^*i*^(*t*) according to the real vaccination rates, which differ from each other in their *I*^*i*^(*t*), which represents the level of close interactions of a vaccinated individual *i* with unvaccinated individuals as a function of time *t* (**Supplementary Methods**).

We simulated different scenarios of infections throughout February 2021. In each date we calculated the observed vaccine effectiveness, 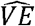, using both the population-based approach (PB) and the secondary infection approach (SI). In the PB-simulated analysis, we performed a simple adjustment to eliminate the heterogeneity in vaccination rates among communities by matching the number of vaccinated and unvaccinated in each community, which can be seen as 1:1 matching. In the SI-simulated analysis we only considered infections within the close-contact circuit, which is similar to a situation of *δ* = 0 in the analytical model (**Supplementary Methods**). Thus, low ⟨*I*⟩ represents a scenario where most close-contacts circuits are partially vaccinated (i.e., a family with only one parent vaccinated), and high ⟨*I*⟩ represents a scenario where most close-contact circuits are either fully vaccinated or not vaccinated at all. In all analyses, we observed clear negative correlations between 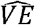 and ⟨*I*⟩, and 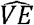 ranged between ~70% and ~95% (**Figure 2A**). We can see that the input *VE* was only obtained for ⟨*I*⟩ at levels around 0.5-0.6 for both the PB and SI simulations. However, the PB-based Dagan et al. study (3) probably had an ⟨*I*⟩ of ~0.25, which is in line with the lockdown that was imposed at this time, while the SI-based Gazit et al. study (4) probably had an ⟨*I*⟩ of ~0.75, which is in line with the limited availability of vaccinations to adults only at this time.

**Figure 2.**
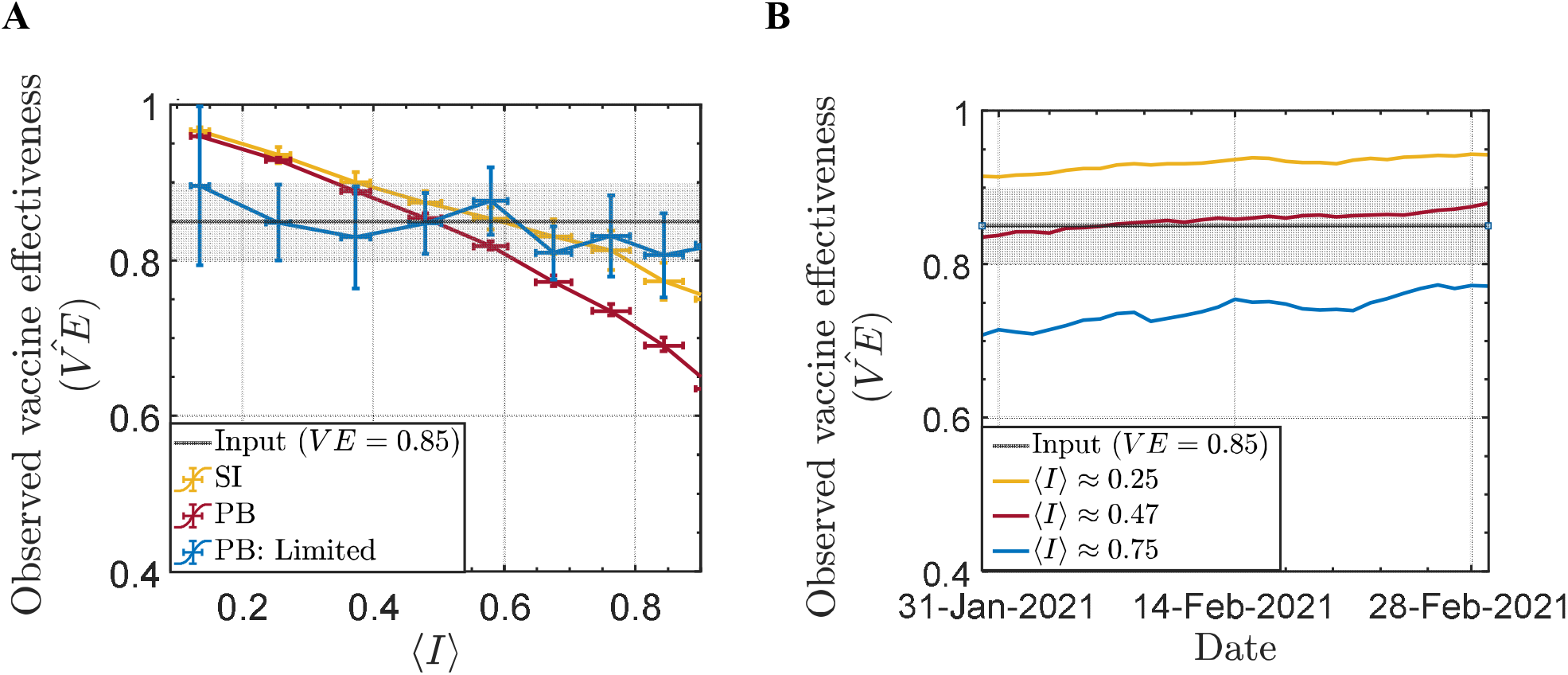
Estimations of 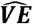 in different levels of mixing between vaccinated and unvaccinated individuals. **A**. Estimations of 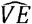 as a function of ⟨**I**⟩. Red line: Population-based (PB) simulations with matching of vaccinated and unvaccinated individuals in each region; Green line: Secondary infection-based (SI) simulations; Blue line: Matched PB with additional exclusion of individuals in close-contact circuits with more than 50% vaccination. **B**. 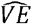 estimations for matched-PB as a function of time. Estimations are shown from January 31 to end of February 28 for three mixing scenarios. For both panels, Grey line – input VE =0.85; Grey band: VE within a 5% uncertainty.

As we can see, even careful matching would be insufficient to retrieve the actual *VE*. This is due to the additional circuits we added in the modelling that aim to mimic household contacts. This creates heterogeneity of ⟨*I*⟩ at the close contacts-level, not just at the community-level. To overcome this issue, we derived an additional analysis that considers the level of mixing. Since it is not simple to adjust for different ⟨*I*⟩ levels in real-world data for PB analysis, we performed the analysis by including only close-contact circuits with over 50% vaccinated individuals. This analysis showed that for PB, it is possible to achieve relatively accurate estimations of *VE* with this relatively simple approach, regardless of ⟨*I*⟩ and time (**Figure 2A** and **Figure S2**). Of note, in SI analysis, both ⟨*I*⟩ and *v* can be theoretically obtained for each family (and *δ* = 0), therefore, 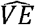 can be adjusted more easily in real-world data (full analysis for 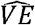 for PB and SI is given in the **Supplementary Methods**).

Finally, we analyzed how 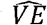 changed over time in the different scenarios (**Figure 2B**). Interestingly, even for ⟨*I*⟩ ≈ 0.5, where the average 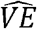 in February was ~85% (PB method; **Figure 2A**), we observed underestimation in early February and overestimation in late February. In general, we noticed that as vaccination rates increased with time, 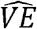 estimations increased accordingly, in line with early assessments of vaccine effectiveness in Israel compared to later assessments. This temporal relationship between the distribution of ⟨*I*⟩ and the vaccination rate adds another layer to the complexity of vaccine effectiveness estimation and can explain divergent results across studies that were performed in different time points.

## DISCUSSION and CONCLUSIONS

We demonstrated here that vaccine effectiveness estimations are sensitive to the dispersion of vaccinations across the population. We found that the level of mixing between vaccinated and unvaccinated individuals can alter the observed vaccine effectiveness. High level of mixing creates indirect protection to unvaccinated individuals and on the other hand increases the risk of infection for vaccinated people, since they frequently interact with unvaccinated people, which in turn leads to underestimation of vaccine effectiveness, while low level of mixing leads to additional protection to the already protected individuals, and in turn to overestimation of vaccine effectiveness. Regardless of what is the real mixing between vaccinated and unvaccinated, which we assume is far from random, it is clear from our analyses that the observed vaccine effectiveness can significantly differ from the real individual-level protection offered by the vaccine.

This study is subject to several limitations. First, this work did not consider that part of the population is protected by previous infection (i.e., recovered). However, the infections data from Israel indicates that by November 2020, a relatively low proportion of the population (no more than 5%) had been infected and recovered. It may affect the assumptions and calculations in this work, but this oversight should result in only a minor error, since we assume that some of the recovered population is also vaccinated. Second, it is not simple to estimate the level of mixing in the population. However, we note that during the time of first estimations of vaccine effectiveness coming from Israel (3), Israel was mostly under a lockdown that was imposed between January 8, 2021, and February 9, 2021. Under the lockdown there were limited interactions overall, which enable us to assume relatively low interaction within the population in Israel and especially low number of interactions between the vaccinated elderly population and the rest of the population which were vaccinated later. It is therefore safe to assume that ⟨*I*⟩ at the time of the study was trending towards 0, or at least much lower than later on. Based on this, we argue that the high vaccine effectiveness reported 95% for adults over 70 years old (3) is substantially overestimated. Furthermore, the lower vaccine effectiveness reported for the 40-69 years old group in that study (90%) is a result of the higher mixing rate of that group. Later, Israel imposed “green passports”, which limit entrance of unvaccinated individuals to crowded places. This is another measure that reduces vaccinated-unvaccinated interactions and may again skew towards overestimation of vaccine effectiveness.

On the other hand, we argue that study designs of secondary household infections underestimate vaccine effectiveness. Gazit et al. used this approach to estimate vaccine effectiveness in Israel and found vaccine effectiveness to be 80% (4). In households, especially in a country with relatively big families as in Israel, most interactions of vaccinated individuals are with unvaccinated children that were not vaccinated at the time of the study. This translates to high ⟨*I*⟩ levels, and as we show this leads to significant underestimation. We note that some of the lower vaccine effectiveness in household infections can be attributed to the prolonged exposure, and in theory, the vaccination is less effective in such scenario. We are aware that the number of family members in Israel is not uniform, however, since the average number of children in Israel is around 3 (26), assuming five people per household is a reasonable assumption.

Our study is not the first to caution about vaccine effectiveness estimations confounded by indirect protection. Previous studies developed a causal inference framework with interference to deal with this issue (19–23). Interestingly, although this issue is well known, in our literature review of vaccine effectiveness studies for COVID-19, we did not identify any study that takes this issue into account. We believe that analytical model provides a novel and relatively simple framework for dealing with vaccine interference, and, as we show here, can be implemented in real-world settings. The analytical model is structured to differentiate between different types of interactions (close and remote), and therefore, better represents real-world infection networks.

In conclusion, we argue here that observed vaccine effectiveness estimations of 90%-95% and 80% stems from a vaccine that probably provides individual-level protection of around 85%. We suggest that by adjusting to the fraction of unvaccinated individuals in a household, in addition to demographic and clinical features, it is possible to reduce the bias when estimating vaccine effectiveness. We hope that future studies will consider the possible bias we highlight in this study and attempt to correct for it, as the rate of effectiveness of the vaccines have high impact on strategies for controlling the spread of the pandemic.

## Data Availability

All data are available online

https://www.cbs.gov.il/EN/Pages/default.aspx

https://data.gov.il/dataset/covid-19.

## ACKNOWLEDGMENTS

DA is supported by the Azrieli Faculty Fellowship and is a Deloro Fellow. We thank Yaniv Erlich, Barak Rave and Michael Geller for fruitful discussions.

