## Supplementary materials for "Over- and under-estimation of vaccine effectiveness"

### Supplementary Methods

#### Analytical heterogenous infection model

The analytical heterogenous infection model is a network composed of multiple cliques, close contact networks, which might resemble household interactions. Nodes in the clique may be 'vaccinated' or 'unvaccinated'. The cliques are weakly connected with each other. We denote $\delta$ as the ratio between infections happening inside the clique versus infections happening outside of the clique. Around half of all infections are household infections (28), so a logical $\delta$ is 1. We also define $\alpha$ as the probability of transmitting the virus; $VE$ as the input for the vaccine effectiveness the reduced risk of infection for protected individuals. Finally, $N_{in}$ is the number of nodes in each clique, and $N_{out}$ is the sum of nodes in all other cliques in the network, where the number of the unvaccinated nodes in all the network, $N_{unvac} = \nu \cdot N_{all} = \nu \cdot (N_{in} + N_{out})$. The parameters used in the HIM are summarized in **Table S1**.

| PARAMETER | INTERPRETATION | VALUE IN HIM |
|---|---|---|
| $\alpha$ | Probability of transmitting the virus | 0.1 (Not needed for $\widehat{VE}$) |
| $VE$ | The input vaccine effectiveness, such that the risk of infection is 1-$VE$ | 0.85 |
| $\delta$ | Infections within outer cliques compared to inner clique infections (normalized to the size of the inner clique) | 1 (half of infections are household infections) |
| $\nu$ | Fraction of unvaccinated people in the total population=$^{N_{unvac}}/_{N_{all}}$ | 0.6 |
| $N_{in}$ | Number of nodes in each clique | 5 |

| | | |
|---|---|---|
| $n$ | Total number of cliques | 5 |
| $N_{all}$ | Total number of nodes | $n \cdot N_{in} = 25$ |

**Table S1**. *Parameters and their interpretation in the heterogenous infection model (HIM).*

We follow (19,21) to further define additional parameters of the number of unvaccinated edges, for each node, inside and outside of the clique. For each node we define a vector $\boldsymbol{A}$, which resemble the number of unvaccinated edges for each node, and later $\boldsymbol{A}$ is used for estimating the amount of interference in the system:

$$\boldsymbol{A^i} = \{A_1^i, A_2^i, A_3^i\} \quad (S.1)$$

**Table S2** summarize these parameters,

| PARAMETER | INTERPRETATION | VALUE |
|---|---|---|
| $A_1^i$ | Binary variable for vaccination for the $i^{th}$ node | 0 – vaccinated<br>1 – vaccinated |
| $A_2^i$ | Number of unvaccinated edges for the $i^{th}$ node in the close clique | |
| $A_3^i$ | Number of unvaccinated edges for the $i^{th}$ node in the remote cliques | for each node:<br>$(1 - A_1^i) + A_2^i + A_3^i = \nu \cdot N_{all}$ |

**Table S2**. *Parameters of vector $A^i$*

The vector $A$ is required for the calculation of the observed vaccine effectiveness ($\widehat{VE}$). Since the aim of this study is to evaluate the effect of unvaccination environment on vaccine effectiveness, for each node, we define $I^i$ as the fraction of edges it has with unvaccinated nodes in the inner clique which will indicate the amount of interference in the system:

$$A_2^i(A_1^i) = I^i(A_1^i) \cdot (N_{in} - 1) \quad (S.2)$$

We further define $\langle I(A_1^i)\rangle$ as the average of all $I^i$ values in the network. A $\langle I(A_i^1 = 1)\rangle = 0$ in the analytical infection model represents a network where the vaccinated and the unvaccinated populations are completely distinct as there are no close interactions between them. As $\langle I(A_i^1 = 1)\rangle$ increases, there is more mixture between the two populations. The observed vaccine effectiveness is defined as:

$$\widehat{VE} = 1 - \frac{\frac{1}{N_{vac}} \sum_{i=1}^{i=N_{vac}} P(A_1^i = 1) =}{\frac{1}{N_{unvac}} \sum_{j=1}^{j=N_{unvac}} P(A_1^j = 1)} \tag{S.3}$$

With $P(A_1^i)$ is individual probability of infection such that $P(A_1^i = 1)$ is the infection probability of the i[th] vaccinated node and $(P(A_1^j = 0)$ is j[th] unvaccinated node. Infection risk from inside the clique depends on the number of vaccinated and unvaccinated nodes in the clique; Infection risk from outside the clique depends on the fraction of vaccinated nodes outside the clique.

$$P = \alpha \times (1 - VE \cdot A_i^1) \tag{S.4}$$

$$\cdot \left\{ \left[(1 - VE) \cdot (N_{in} - 1 - A_2^i) + A_2^i\right] + \delta \frac{N_{in} - 1}{N^{out}} \cdot \left[(1 - VE) \cdot (N_{out} - A_3^i) + A_3^i\right] \right\}$$

$$P^i = \alpha \times (1 - VE \cdot A_i^1)$$

$$\cdot \left\{ (1 - VE) \cdot (N_{in} - 1) + A_2^i \cdot VE \ + \delta \frac{N_{in} - 1}{N_{out}} \cdot \left[(1 - VE) \cdot N_{out} + A_3^i \cdot VE\right] \right\} =$$

$$\alpha \times (1 - VE \cdot A_i^1)$$

$$\cdot \left\{ (1 - VE) \cdot (N_{in} - 1) + A_2^i \cdot VE \ + \delta (N_{in} - 1) \cdot (1 - VE) + \delta \frac{N_{in} - 1}{N_{out}} \cdot A_3^i \cdot VE \right\}$$

$$\alpha \times (1 - VE \cdot A_i^1) \cdot \left\{ (1 - VE) \cdot (N_{in} - 1) \cdot (1 + \delta) + VE \cdot [A_2^i \ + \delta \frac{N_{in} - 1}{N_{out}} \cdot A_3^i] \right\}$$

Where:

$$A_3^i = \nu \cdot N_{all} - (1 - A_1^i + A_2^i) \tag{S.5}$$

So, eq. S.2 becomes:

$$P = \alpha \times (1 - VE \cdot A_i^1)$$

$$\cdot \left\{ (1 - VE) \cdot (N_{in} - 1) \cdot (1 + \delta) + VE \cdot \left[ A_2^i \ + \delta \frac{N_{in} - 1}{N_{out}} \cdot (\nu \cdot N_{all} - 1 - A_2^i + A_1^i) \right] \right\} \tag{S.6}$$

Since only $A_1^i$ and $A_2^i$ depend on $I$, we divided eq S.4 into two parts: F1 which is a function of $A_1^i$ and F2 which is a function of $A_1^i$ and $A_2^i$:

$$P = \alpha \times \left(1 - VE \cdot A_i^1\right) \cdot \left\{(1 - VE) \cdot (N_{in} - 1) \cdot (1 + \delta) + VE \cdot \left[\delta \frac{N_{in} - 1}{N^{out}} \cdot \left(\nu \cdot N^{all} - 1 + A_1^i\right)\right]\right\} + \alpha \times \left(1 - VE \cdot A_i^1\right) \cdot \left\{VE \cdot A_2^i \left[1 - \delta \frac{N_{in} - 1}{N^{out}}\right]\right\} \quad (S.7)$$

With:

$$F1 = \alpha \times \left(1 - VE \cdot A_1^i\right) \cdot \left\{(1 - VE) \cdot (N_{in} - 1) \cdot (1 + \delta) + VE \cdot \left[\delta \frac{N_{in} - 1}{N^{out}} \cdot \left(\nu \cdot N_{in} + A_1^i - 1\right)\right]\right\} \quad (S.8)$$

$$F2 = \alpha \times \left(1 - VE \cdot A_1^i\right) \cdot VE \cdot A_2^i \cdot \left(1 - \delta \frac{N_{in} - 1}{N_{out}}\right) \quad (S.9)$$

And

$$A_2^i = I^i(A_1^i) \cdot (N_{in} - 1) \quad (S.10)$$

For the vaccinated nodes:

$$\left\langle P\left(A_1^i = 1\right)\right\rangle = \frac{1}{N_{vac}} \sum P\left(A_1^i = 1\right) = \frac{1}{N_{vac}} \sum F1\left(A_1^i = 1\right) + \frac{1}{N_{vac}} \sum F2\left(A_1^i = 1\right) =$$
$$\frac{1}{N_{vac}} \sum \alpha \times (1 - VE) \cdot \left\{(1 - VE) \cdot (N_{in} - 1) \cdot (1 + \delta) + VE \cdot \left(\delta \frac{N_{in} - 1}{N_{out}} \cdot \nu \cdot N_{all}\right)\right\}$$
$$+ \frac{1}{N_{vac}} \sum \alpha \times (1 - VE) \cdot \left\{VE \cdot \left[\boldsymbol{I}^i\left(A_1^i = 1\right) \cdot (N_{in} - 1)\right] \cdot \left(1 - \delta \frac{N_{in} - 1}{N_{out}}\right)\right\} =$$
$$\alpha \times (1 - VE) \cdot \left\{(1 - VE) \cdot (N_{in} - 1) \cdot (1 + \delta) + VE \cdot \left[\delta \frac{N_{in} - 1}{N_{out}} \cdot (\nu \cdot N_{all})\right]\right\} +$$
$$\alpha \times (1 - VE) \cdot \left\{VE \cdot \left[\left\langle I\left(A_1^i = 1\right)\right\rangle \cdot (N_{in} - 1)\right] \cdot \left(1 - \delta \frac{N_{in} - 1}{N_{out}}\right)\right\} \quad (S.11)$$

For the unvaccinated nodes:

$$\left\langle P\left(A_1^j=0\right)\right\rangle = \frac{1}{N_{unvac}}\sum P\left(A_1^j=1\right) = \frac{1}{N_{vac}}\sum F1\left(A_1^j=0\right) + \frac{1}{N_{vac}}\sum F2\left(A_1^j=0\right) =$$

$$\frac{1}{N_{unvac}}\sum\alpha\times\left\{(N_{in}-1)\cdot(1+\delta)+VE\cdot\left[\delta\frac{N_{in}-1}{N_{out}}\cdot(\nu\cdot N_{all}-1)\right]\right\}$$

$$+\frac{1}{N_{unvac}}\sum\alpha\times\left\{VE\cdot\left[I\left(A_1^i=0\right)\cdot(N_{in}-1)\right]\cdot\left(1-\delta\frac{N_{in}-1}{N^{out}}\right)\right\} = \qquad (S.12)$$

$$\alpha\times\left\{\left(N^{in}-1\right)\cdot(1+\delta)+VE\cdot\left[\delta\frac{N_{in}-1}{N_{out}}\cdot(\nu\cdot N_{all}-1)\right]\right\}+$$

$$\alpha\times\left\{VE\cdot\left[\left\langle I\left(A_1^j=0\right)\right\rangle\cdot(N_{all}-1)\right]\cdot\left(1-\delta\frac{N_{in}-1}{N_{out}}\right)\right\}$$

We can see that $\langle I\left(A_1^i=0\right)\rangle$ can be written in the following form:

$$\langle I\left(A_1^i=0\right)\rangle = 1-\tfrac{1-\nu}{\nu}\left\langle I(A_1^j=1\right\rangle, \qquad (S.13)$$

and Eq S.10 becomes:

$$\left\langle P\left(A_1^j=0\right)\right\rangle = \alpha\times\left\{(N_{in}-1)\cdot(1+\delta)+VE\cdot\left[\delta\frac{N_{in}-1}{N_{out}}\cdot(\nu\cdot N_{all}-1)\right]\right\}+ \qquad (S.14)$$

$$\alpha\times\left\{VE\cdot\left[1-\frac{1-\nu}{\nu}\langle I\left(A_1^i=1\right)\rangle\right]\cdot(N_{in}-1)\cdot\left(1-\delta\frac{N_{in}-1}{N_{out}}\right)\right\}$$

We denote $\left\langle I(A_1^i=1\right\rangle$ as $\langle I\rangle$, as the average number of close vaccinated-unvaccinated interactions per for the vaccinated nodes, and $\delta' = \delta\,\frac{N_{in}-1}{N_{out}}$ and $N'_{in}=N_{in}-1$, Eq (S.3) becomes:

$$\widehat{VE} = 1-\frac{\frac{1}{n_{vac}}\sum P\left(A_1^i=1\right)}{\frac{1}{n_{unvac}}\sum P\left(A_1^i=0\right)} = \qquad (S.15)$$

$$VE\times\frac{N'_{in}\cdot(1+\delta)+VE\cdot\delta'\cdot\nu\cdot N^{all}+VE\cdot\langle I\rangle\cdot N'_{in}\cdot(1-\delta')}{N'_{in}\cdot(1+\delta)+VE\cdot\delta'\cdot(\nu\cdot N^{all}-1)+VE\cdot\left[1+\langle I\rangle\left(1-\frac{1}{\nu}\right)\right]\cdot N'_{in}\cdot(1-\delta')}$$

Note that $\widehat{VE}$ is independent of $\alpha$.

We wish to find the conditions for which Eq. (S.15) is not equal to $VE$**.** Since $\delta' < 1$ $\nu < 1$ and $\langle I \rangle \leq \frac{N_{in}}{N_{in}-1}\nu$ the 1 denominator of Eq. (S.15) is always positive so the sign of the term $\frac{VE\cdot\left[\delta\prime - \frac{N\prime_{in}\,(\delta'-1)(\langle I\rangle-\nu)}{\nu}\right]}{N'_{in}\cdot(1+\delta)+VE\cdot\delta'\cdot\left(\nu\cdot N^{all}-1\right)+VE\cdot\left[1+\langle I\rangle\left(1-\frac{1}{\nu}\right)\right]\cdot N'_{in}\cdot\,(1-\delta')}$ determined by the sign of the numerator. Similar to (19,21),20]. we can now define the average total causal effect on vaccine effectiveness as:

$$\overline{TVE}\left(\langle I\rangle, \delta, \nu, N^{in}, N^{all}\right) = \widehat{VE}(\langle I\rangle, \delta, \nu, N_{in}, N_{all}) - VE \qquad \textit{(S.17)}$$

Such that:

$$\begin{aligned}
&\overline{TVE}(\langle I\rangle, \delta, \nu, N_{in}, N_{all}) \geq 0 \quad \langle I\rangle \leq \nu\left[1 - \frac{\delta'}{N'_{in}(1-\delta')}\right] \\
&\overline{TVE}(\langle I\rangle, \delta, \nu, N_{in}, N_{all}) < 0 \quad \langle I\rangle > \nu\left[1 - \frac{\delta'}{N'_{in}(1-\delta')}\right]
\end{aligned} \qquad \textit{(S.18)}$$

Interestingly, for $\delta = 0$, i.e, only household infection, we find that:

$$\begin{aligned}
&\overline{TVE}(\langle I\rangle, \delta, \nu, N_{in}, N_{all}) \geq 0 \quad \langle I\rangle \leq \nu \\
&\overline{TVE}(\langle I\rangle, \delta, \nu, N_{in}, N_{all}) < 0 \quad \langle I\rangle > \nu
\end{aligned} \qquad \textit{(S.19)}$$

**MAM: Flexible Monte-Carlo Agent based Model for Modelling COVID-19 Spread**

In this work, we are using MAM, a flexible Monte-Carlo Agent based Model for modelling COVID-19 spread (24), originally introduced by De-Leon and Pederiva (29,30) to model the spread of COVID-19 in Israel in the presence of effective vaccines from December 2020 until March 2021, and as a result, to estimate the observed vaccine effectiveness. As opposed to other infection models, such as the Susceptible Infected Removed (SIR) (31–33), this particle model enables us to distinguish between different age groups and treat each one separately, assuming that the infection occurs throughout the population simultaneously. Furthermore, a particle model can be adjusted to the actual rate of population vaccination. This model enables us to accurately examine the different effects of the vaccine on subgroups of the vaccinated population and the entire population. We used numerical simulations that consist of $\mathbf{9.2 \cdot 10^6}$ particles (which simulates the number of residents in Israel), where each particle has a number from 1 to $\mathbf{9.2 \cdot 10^6}$ under the assumption of three infection circuits (arranged according to the likelihood of infection from high to low): a household infection cycle involving five people (which is the average in Israel); community based-infection, an infection cycle of 25 people; and infections in a remote community – an infection circle of 125 people. We define the model to generate half of all infections in the simulation to occur within households, by assuming that particle 1 interacts mostly with particles 2-5 (i.e., particles 1-5 resemble a household, so any group of 5 particles). Still, there is almost no contact between 1 and particle number 2000.

**Modelling the spread of COVID-19 in the presence of effective vaccines**

For modeling the spread of a pandemic, the most essential input required to define is $R$, the reproduction rate; when $R$ is above 1, one individual infects, on average, more than one other individual, which indicates the disease is spreading. We distinguish between $R_0$, the basic reproduction number; and $R_t$, the theoretical reproduction rate of the disease. $R_t$ is an estimation of the rate of encounters between infected and non-infected individuals that would have resulted in an infection without vaccinations. In MAM, $R_t$ is the particle density or the size of the area for the particles, which is similar for all age groups. In this work, following the easing of social restrictions in Israel in February 2021, we estimate the theoretical $R_t$ in Israel from January 2021 until February 2021 to be 1.2 (34). Another inputs required for modeling the spread of COVID-19 in the presence of effective vaccine is the effectiveness of the vaccine ($VE$). We assumed the protection from the vaccine starts seven days after the first dose and reached its maximum after seven days from the second dose (1,3,24). Also, the vaccination rate of the population is required, as detailed in the next subsection.

**Adjustment for real-world data**

We use publicly available data from Israel (25) for 1,578 different statistical areas, which are communities defined by the Central Bureau of Statistics of Israel and include communities of about 5,000 individuals on average (26). Using this data allows us high resolution of the heterogeneity in vaccinations uptake. We use the community-level vaccination data between the end of December 2020 and March 2021 to calculate the daily number of confirmed cases in Israel for each statistical area. We assume that most infections are local, and half of the infections occur at home, and only less than 1% of the infections occur outside the statistical area.

For modeling the spread of COVID-19 using real-world data, we create for each particle a vector $A^i$, similar to that introduced for the analytical infection model. However, since this is a time-dependent model, for that case $A^i \rightarrow A^i(t)$, which the rate of vaccination is determined by the real vaccination rate in Israel. For each community, the number of people vaccinated each day is determined by the actual vaccination rates. Still, we can choose who will be vaccinated that day from that area. As a result, we developed ten different vaccination scenarios, which differ in the degree of mixing between vaccinated and non-vaccinated over time. Hence, for each scenario, we created a different array of $A^i(t)$ according to the real vaccination rate, which differ from each other by $I(t)$, which is one of the inputs needed for simulation (see (24)) which affect the number of interactions between vaccinated and unvaccinated (differentI $\langle I \rangle$ levels). Consequently, the vaccination order within each community is important since the greatest chance of infection is between two adjacent serial numbers. Since not all populations were vaccinated simultaneously in reality, $\langle I \rangle = \langle I(t) \rangle$ . As a result, we define for every day a mean value of $I$, $\langle I(t) \rangle$, which represents the daily average percentage of vaccined-unvaccinated interactions for the vaccinated population. Note that we assume that half of all infections occur in the first infection circuit (at home). Therefore, the contribution from the second and third circuits is lower than the contribution from the first circuit.

All the data and code used in these analyses are available on: https://github.com/hdeleon1/Over-and-under-estimation-

##### Calculation of the observed vaccine effectiveness

For calculating vaccine effectiveness, we defined $\widehat{VE}$, seven days after the second dose for each day as:

$$\widehat{VE}(t) = 1 - \frac{CC_{vac}(t)/All_{vac}(t)}{CC_{unvac}(t)/All_{unvac}(t)} \quad (4)$$

where:

- $CC_{vac}(t)$ - daily number of particles who became infected seven days or more after receiving the second dose of the vaccine.
- $CC_{non}(t)$ - daily number of particles who became infected before receiving the first dose of the vaccine.
- $All_{vac}(t)$ - daily number of fully vaccinated particles in the population (seven days or more from receiving the second dose of the vaccine).
- $All_{non}(t)$ - daily number of unvaccinated people in the population (before receiving the first dose of the vaccine).

We used two methods for calculating the daily vaccine effectiveness for each vaccination scenarios using two different methods (**Figure 2**). The crude approach, which is just counting daily infections, and the matched approach, where similar amount of vaccinated and unvaccinated individuals are chosen randomly from each statistical area. Matching across statistical areas reduces the effect of the heterogeneity in vacciations uptake, and in theory should eliminate the overestimation caused by this. However, since our model consists of a ’family’ circuit, the matching is insufficient for avoiding the overestimation. We note that this is most probably true. For example, if one partner is vaccinated, most likely, the other partner will be vaccinated as well.

**Full analysis of the vaccine effectiveness:**

In the main result section, we presented vaccine effectiveness estimations for three cases (SI, PB, and PB+limited). Here we present the full analysis for each method (PB and SI). We used three approaches in PB and SI for the calculations: crude analysis, matching and matching + families.

The scenarios simulated infections throughout February 2021. In each date we calculated the observed vaccine effectiveness, $\widehat{VE}$ using the population-based approach (PB) and the secondary infection approach (SI)**.** The analysis showed a clear negative correlation between $\widehat{VE}$ and $\langle I \rangle$ for both the PB and SI analysis (**Figure S2A-B**). Interestingly, in the PB analysis we observed that for all the range of $\langle I \rangle, \widehat{VE}$ was higher than $VE$, which was 85%. The explanation for this result is that there was high heterogeneity in vaccination uptake across communities in Israel (**Figure S1**). While most communities have reached high vaccination rates rapidly, there were some communities with low vaccination uptake, leading to a bimodal distribution of vaccinations rates, and in turn of the distribution of $\langle I \rangle$ (**Figure S2C**). In those low vaccinated communities, there is low indirect protection, and the heterogeneity across the population is what is causing the high overestimation across the whole range of $\langle I \rangle$. This result should warrant that crude vaccine effectiveness estimations in heterogenous population are bound to overestimate vaccine effectiveness.

When using the SI approach for calculating $\widehat{VE}$, we observed an even stronger negative correlation between $\widehat{VE}$ and $\langle I \rangle$. In this analysis we only consider infections within the close-contact circuit, which is similar to a situation of $\delta = 0$ in analytical model (Eq S.19). Thus, low $\langle I \rangle$ represents a scenario where most close-contacts circuits are partially vaccinated (i.e., a family with only one parent vaccinated), and high $\langle I \rangle$ represents a scenario where most close-contact circuits are either fully vaccinated or not vaccinated at all. Similar to the analytical analysis we see in our real-world modeling that if $\langle I \rangle > 0.5$ , the observed $\widehat{VE}$ is lower than $VE$.

We next performed the same analysis, but this time $\widehat{VE}$ was calculated by matching the number of vaccinated and unvaccinated in each community, which eliminates the heterogeneity in vaccination rates among communities, and is similar to a matched analysis of $\widehat{VE}$, where individual-level data is available (**Figure S2A-B**). After matching for both SI and PB analyses, $\widehat{VE}$ could still be both under- and overestimated. The estimations of $\widehat{VE}$ in the matched analysis was from ~60% to 95%, depending on $\langle I \rangle$. $VE$ was only obtained for $\langle I \rangle$ at levels around 0.5 and 0.3 for PB and SI, respectively. The reason why matching is still insufficient to retrieve $VE$ stems from the additional circuits we added in the modelling that aim to mimic household contacts. This creates heterogeneity of $\langle I \rangle$ at the close contacts-level, not just at the community-level. To overcome this issue, we derived an additional analysis that considers the level of mixing. Since it is not simple to adjust for different $\langle I \rangle$ levels in real-world data for PB analysis, we performed the analysis by include only close-contact circuits with over 50% vaccinated individuals. This analysis showed that for PB, it is possible to achieve relatively accurate estimations of $VE$ with this relatively simple approach. Of note, in SI analysis, both $\langle I \rangle$ and $\nu$ can be theoretically obtained for each family (and $\delta = 0$) , therefore, $\widehat{VE}$ can be adjusted more easily in real-world data.

The heterogeneity of $\langle I \rangle$ is also a function of time. As vaccination rates increase, the distribution of $I$ changes. A possible real-world scenario that illustrates this is when vaccinations were provided for 12-17 year old children. In families with children in those ages, the $\langle I \rangle$ values were reduced, while $\langle I \rangle$ did not change in families with younger children.

#### Heterogeneity in vaccination uptake in Israel

The vaccination campaigns started by vaccinating older individuals before moving to younger populations. Further, there is a significant association between vaccine uptake and socioeconomic status (SES) and other factors (35). In Israel, based on the data from (26) for the population of each statistical area and the data from (25) for the daily A vaccinated people for each of the 1,578 statistical areas, we can calculate, on a daily basis what is the percentage of the population which is vaccinated with two doses of vaccine. We found that the distribution of the percentage of the population that was vaccinated with two vaccine doses on April 1, 2021, manifested in a somewhat bi-modal distribution of vaccine uptake across the population: while in the majority of the statistical regions (>90%), we observe a normal distribution around 60% with standard deviation of 10% 4 months after the beginning of the campaign, in 10% of regions, the vaccination rate was only <25% (**Figure S2A**). This heterogeneity in vaccine uptake is even more pronounced in the first 45 days of the vaccination campaign.

### Supplementary Figure 1

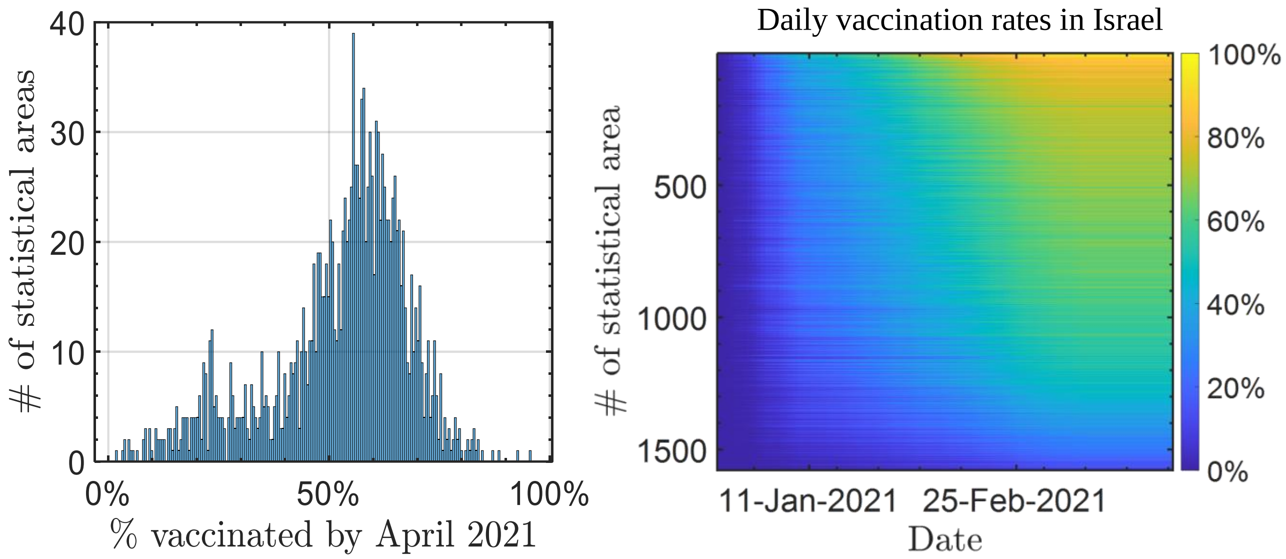


**Figure S1.** ***Distribution of vaccination in Israel. Vaccine uptake for COVID-19 in Israel is not uniform across the population. A.*** *Distribution of the rate of fully vaccinated individuals in 1,578 communities in Israel on April 1, 2021.* ***B.*** *Daily vaccination rates in Israel for each of the 1,578 statistical. Rows represent statistical communities, columns represent days, and the color is the cumulative percentage of vaccinated individual in the community.*

#### Supplementary Figure 2

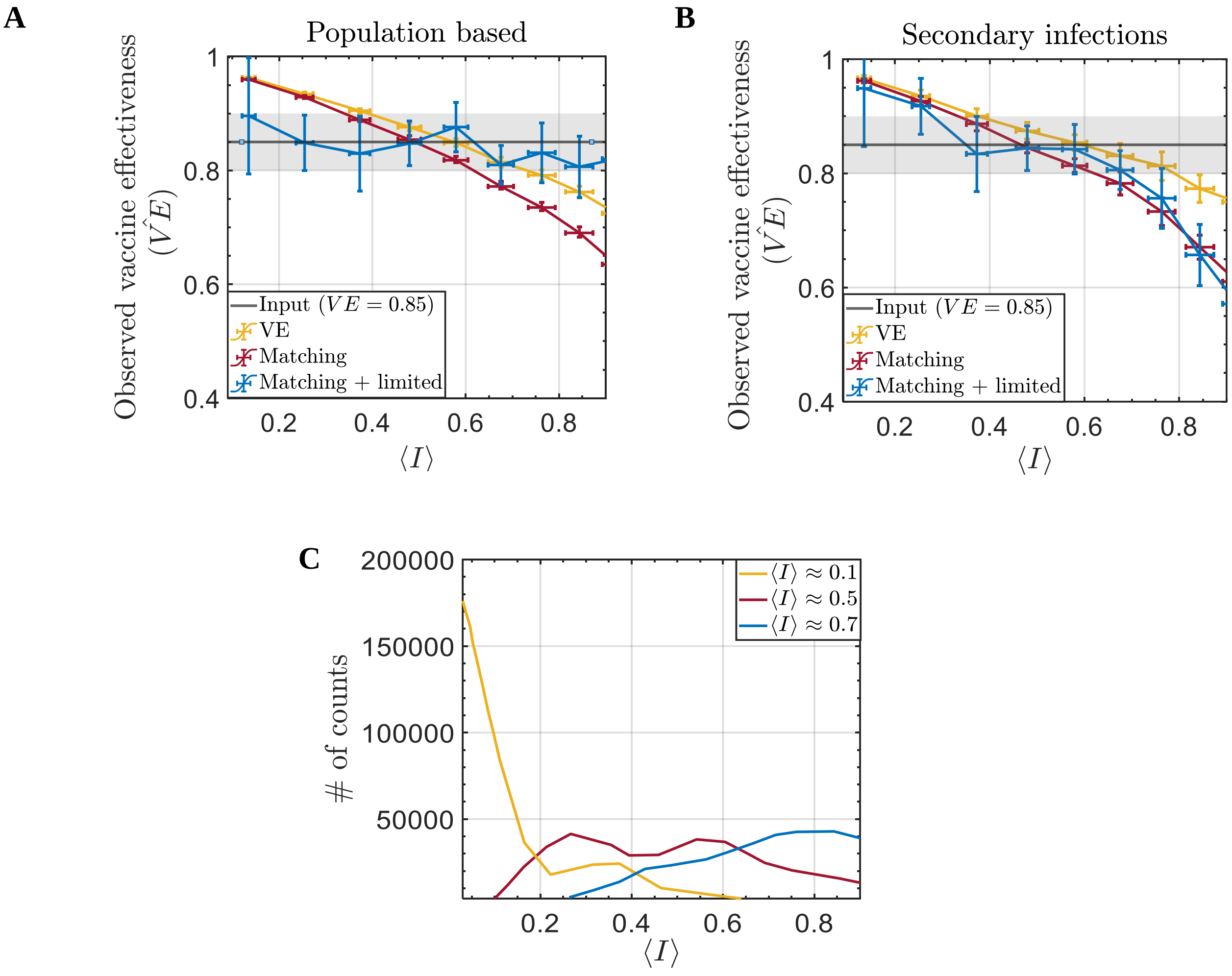


***Figure S2. A.*** *Estimations of $\widehat{VE}$ as a function of $\langle \boldsymbol{I} \rangle$ using population-based analysis. Yellow line: crude analysis; red line: with matching of vaccinated and unvaccinated individuals in each statistical area; blue line: matching for statistical areas and additionally filtering out individuals in close-contact circuits with more than 50% vaccination. Grey line: input VE =0.85; Grey band: VE within a 5% uncertainty.* ***B.*** *Similar to A, but for secondary infection-based analysis.* ***C.*** *Distribution of $\langle I \rangle$ in all vaccinated individuals on February 1, 2021, in three scenarios of mixing vaccinations across the population.*

#### Supplementary Table 3

| | Nodes No. | Binary variable for vaccination, $A_1^i$ | | | No. of edges with unvaccinated nodes in inner clique, $A_2^i$ | | | No. of edges with unvaccinated nodes in remote clique, $A_3^i$ | | | $I(A_1^i)$ | | |
|---|---|---|---|---|---|---|---|---|---|---|---|---|---|
| Network No. | | A | B | C | A | B | C | A | B | C | A | B | C |
| Clique 1 | 1 | 1 | 0 | 0 | 0 | 0 | 2 | 15 | 14 | 13 | 0 | 0 | 0.5 |
| | 2 | 1 | 1 | 0 | 0 | 1 | 2 | 15 | 14 | 13 | 0 | 0.25 | 0.5 |
| | 3 | 1 | 1 | 0 | 0 | 1 | 2 | 15 | 14 | 13 | 0 | 0.25 | 0.5 |
| | 4 | 1 | 1 | 1 | 0 | 1 | 3 | 15 | 14 | 13 | 0 | 0.25 | 0.75 |
| | 5 | 1 | 1 | 1 | 0 | 1 | 3 | 15 | 14 | 13 | 0 | 0.25 | 0.75 |
| Clique 2 | 6 | 1 | 0 | 0 | 0 | 1 | 2 | 15 | 13 | 13 | 0 | 0.25 | 0.5 |
| | 7 | 1 | 0 | 0 | 0 | 1 | 2 | 15 | 13 | 13 | 0 | 0.25 | 0.5 |
| | 8 | 1 | 1 | 1 | 0 | 2 | 3 | 15 | 13 | 13 | 0 | 0.5 | 0.75 |
| | 9 | 1 | 1 | 1 | 0 | 2 | 3 | 15 | 13 | 13 | 0 | 0.5 | 0.75 |
| | 10 | 1 | 1 | 0 | 0 | 2 | 2 | 15 | 13 | 13 | 0 | 0.5 | 0.5 |
| Clique 3 | 11 | 0 | 1 | 1 | 4 | 3 | 3 | 10 | 12 | 13 | 1 | 0.75 | 0.75 |
| | 12 | 0 | 1 | 1 | 4 | 3 | 3 | 10 | 12 | 13 | 1 | 0.75 | 0.75 |
| | 13 | 0 | 0 | 0 | 4 | 2 | 2 | 10 | 12 | 13 | 1 | 0.5 | 0.5 |
| | 14 | 0 | 0 | 0 | 4 | 2 | 2 | 10 | 12 | 13 | 1 | 0.5 | 0.5 |
| | 15 | 0 | 0 | 0 | 4 | 2 | 2 | 10 | 12 | 13 | 1 | 0.5 | 0.5 |
| Clique 4 | 16 | 0 | 0 | 1 | 4 | 3 | 3 | 10 | 11 | 13 | 1 | 0.75 | 0.75 |
| | 17 | 0 | 1 | 0 | 4 | 4 | 2 | 10 | 11 | 13 | 1 | 1 | 0.5 |
| | 18 | 0 | 0 | 0 | 4 | 3 | 2 | 10 | 11 | 13 | 1 | 0.75 | 0.5 |
| | 19 | 0 | 0 | 0 | 4 | 3 | 2 | 10 | 11 | 13 | 1 | 0.75 | 0.5 |
| | 20 | 0 | 0 | 1 | 4 | 3 | 3 | 10 | 11 | 13 | 1 | 0.75 | 0.75 |
| Clique 5 | 21 | 0 | 0 | 1 | 4 | 4 | 3 | 10 | 10 | 13 | 1 | 1 | 0.75 |
| | 22 | 0 | 0 | 0 | 4 | 4 | 2 | 10 | 10 | 13 | 1 | 1 | 0.5 |
| | 23 | 0 | 0 | 0 | 4 | 4 | 2 | 10 | 10 | 13 | 1 | 1 | 0.5 |
| | 24 | 0 | 0 | 0 | 4 | 4 | 2 | 10 | 10 | 13 | 1 | 1 | 0.5 |
| | 25 | 0 | 0 | 1 | 4 | 4 | 3 | 10 | 10 | 13 | 1 | 1 | 0.75 |

**Table S3.** *The values of vector $A^i$, for each node, in each of the networks on figures 1 A-C.*
